# Tracking Antimicrobial Resistant Organisms Timely (TAROT): A Workflow Validation Study for Successive Core-genome SNP-based Nosocomial Transmission Analysis

**DOI:** 10.1101/2025.01.23.25320873

**Authors:** Kotaro Aoki, Kohji Komori, Tetsuo Yamaguchi, Sohei Harada, Mayumi Tsukada, Hinako Murakami, Kazuhiro Tateda

## Abstract

**Background:** Effective infection prevention and control (IPC) interventions in hospitals require timely information to determine the potential transmission of antimicrobial-resistant (AMR) organisms.

**Objectives:** We validated a successive core-genome single nucleotide polymorphism (cgSNP)-based phylogenetic analysis workflow, ‘Tracking Antimicrobial Resistant Organisms Timely’ (TAROT), with Oxford Nanopore Technologies (ONT) sequencer for methicillin-resistant *Staphylococcus aureus* (MRSA).

**Methods:** We have developed a TAROT workflow for successive phylogenetic analysis using ONT data. We sequenced 34 MRSA strains isolated from Toho University Omori Medical Center using MinION (ONT) and MiSeq (Illumina). Each strain’s ONT data were inputted into TAROT (TAROT-ONT), and successive cgSNP-based phylogenetic analyses were conducted. Illumina data were processed with a batched cgSNP-based phylogenetic analysis. Assembly-based analysis identified AMR genes, AMR mutations, and virulence genes.

**Results:** MinION generated an average sequence depth of 262x for the ST8 reference genome within three hours. TAROT-ONT successively generated 11 phylogenetic trees for 14 ST8 strains, seven trees for 10 ST1 strains, and two trees for five ST5 strains. Highly suspected transmission pairs (pairwise cgSNP < 5) were detected in trees #6 through #11 for ST8, trees #3, #5, and #7 for ST1, and tree #2 for ST5. Differences in cgSNP value between TAROT-ONT and Illumina ranged from zero to two within pairs with fewer than 20 cgSNPs using Illumina. TAROT-ONT bioinformatic analysis for each strain required five to 42 minutes. The identification of AMR genes, mutations, and virulence genes showed high concordance between ONT and Illumina.

**Conclusion:** TAROT-ONT facilitates effective IPC intervention for MRSA nosocomial transmissions by providing timely feedback through cgSNP-based phylogenetic analyses.

## Introduction

The transmission of antimicrobial-resistant (AMR) organisms in hospitals is a critical issue that undermines patient safety and results in substantial economic losses.^1–3^ The rising number of AMR organisms identified in clinical microbiology laboratories may signal their spread within hospitals. Among these, methicillin-resistant *Staphylococcus aureus* (MRSA) is closely monitored in most hospitals, and a short-term increase in the MRSA isolation rate may indicate active transmission.^4^ MRSA clones can frequently spread within hospitals, even when patients carrying MRSA are quarantined and contact precautions are implemented.^5^ Effective prevention of nosocomial transmission requires detailed information to verify whether clonal transmission of MRSA strains is occurring, but this cannot be determined through routine microbiological testing.^6–8^

Whole-genome sequencing (WGS) allows for single nucleotide polymorphism (SNP) analysis within the core-genome (cgSNP analysis), which is a gold-standard method for quantitatively determining genetic relationships among strains, replacing traditional methods such as pulsed-field gel electrophoresis and multilocus variable-number tandem repeat analysis.^7,9–15^ cgSNP analysis was first applied to the investigation of MRSA outbreaks.^16^ The method quantitatively evaluates the genetic relationships between strains based on the number of detected cgSNPs. The thresholds of cgSNPs that support short-term nosocomial transmission depend on the evolutionary rate and within-host variation of the species under consideration. Coll et al. proposed a conservative cgSNP threshold for MRSA, suggesting that transmission within six months can be ruled out if more than 15 cgSNPs are identified.^17^

Using genome epidemiology of AMR organisms with WGS and cgSNP analysis in healthcare settings provides crucial insights to enhance and target infection prevention and control (IPC) interventions. The routine implementation of weekly WGS for MRSA not only improves the accuracy of outbreak detection but also enables the de-escalation of IPC activities by identifying pseudo-outbreaks.^18^ However, frequent WGS and timely reporting to the IPC team would have an even greater impact on IPC decision-making. To date, cgSNP analysis has been conducted using short-read data generated by high-throughput sequencers (HTS), specifically the Illumina system, to reduce costs through batch sequencing.^19–21^ However, batch sequencing is time-consuming as it requires the collection of a large number of samples for a single analysis, making it challenging to provide cgSNP analysis results promptly and timely enough to influence IPC intervention.^22,23^ MinION/Flongle (Oxford Nanopore Technologies, ONT, Oxford, UK) offers a cost-effective and scalable solution for daily, single-strain WGS.^24–27^ Nevertheless, most automated bioinformatic tools and workflows for cgSNP analysis and genome epidemiology are optimized for short-read HTS data.^28,29^

To close the gap between ONT sequencing and automated bioinformatic workflows for efficient IPC interventions, we propose ‘Tracking Antimicrobial Resistant Organisms Timely’ (TAROT)(Figure 1). TAROT facilitates automated, successive cgSNP-based phylogenetic analyses by using ONT data on the server side, eliminating the need for high-performance machines on the client side, such as in clinical microbiology laboratories. TAROT includes databases that accumulate core genome data, categorize it by strain lineage, and conduct cgSNP-based phylogenetic analysis based on the input of each strain lineage. In this study, we validated the performance of cgSNP analysis using ONT data within TAROT (TAROT-ONT) by comparing it to cgSNP analysis based on Illumina data, applied to MRSA strains, which serve as a model AMR pathogen.

**Figure 1.**
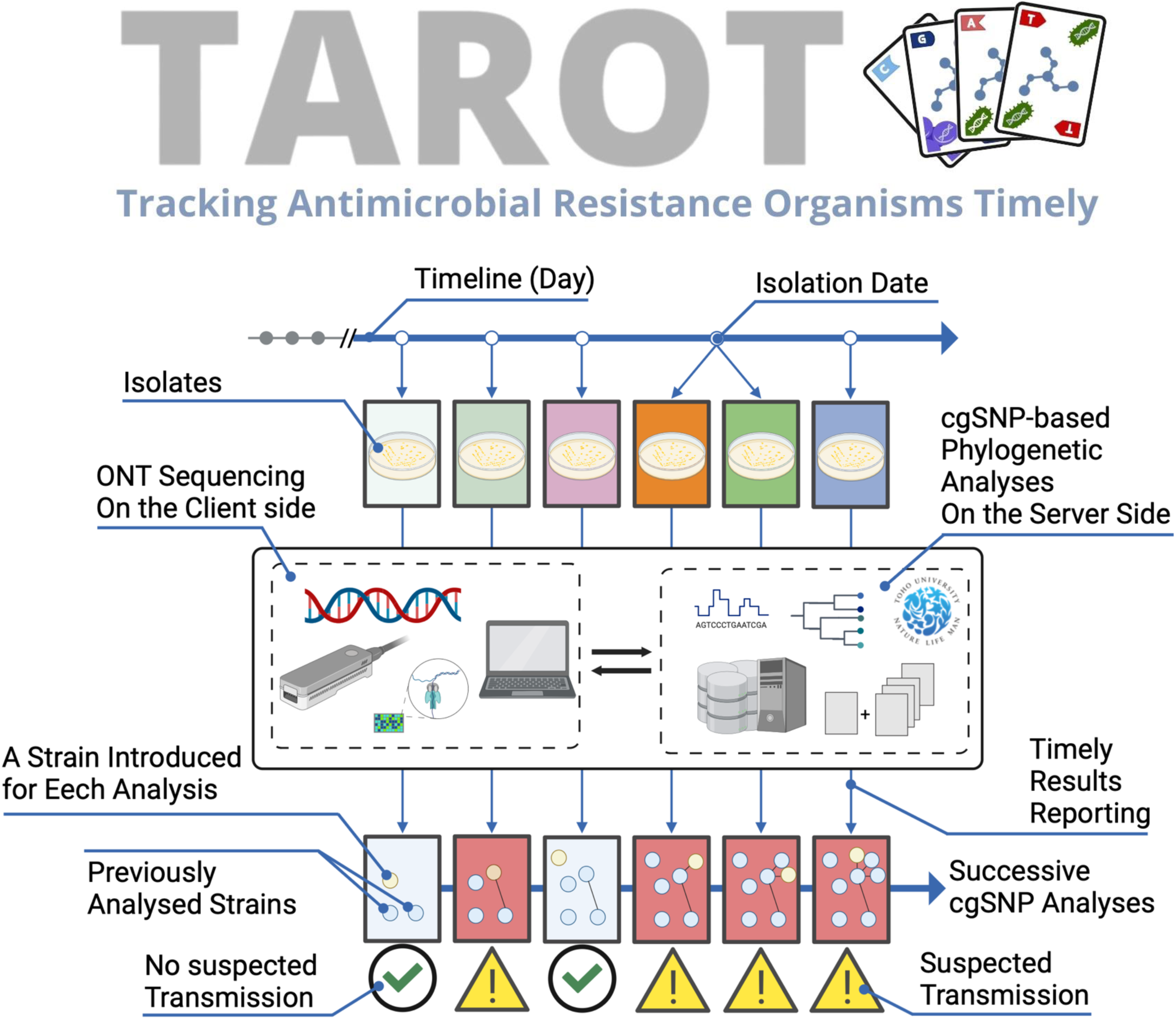
Concept of Tracking Antimicrobial Resistance Organisms Timely (TAROT) TAROT provides on-demand, successive analyses of core-genome single nucleotide polymorphisms (cgSNP). On the client side, each isolate is sequenced using an Oxford Nanopore Technologies sequencer. The resulting sequence data are then transferred to and analyzed on the server side, and the cgSNP analysis results are reported for each isolate. Timely feedback can help enhance infection prevention and control interventions in hospitals.

## Materials and Methods

### Strains

Among the MRSA strains isolated from patients and ward environments at Toho University Omori Medical Center for infection control investigations between May 2011 and February 2022, 34 strains were used in this study. Before planning this study, a draft WGS using MiSeq (Illumina Inc., CA, USA) and cgSNP-based phylogenetic analysis had been conducted to inform IPC interventions. The 34 MRSA strains used in this study were arbitrarily selected based on their suitability for validating TAROT, referencing the previously conducted cgSNP-based phylogenetic analysis with MiSeq data. Some strains were selected from the three frequently collected STs (ST8, ST1, and ST5) to assess whether TAROT could identify the number of cgSNPs needed to distinguish strains within the same ST for suspected transmission. Other strains were selected from different STs to verify the functionality of the automated database classification systems across different STs. In total, the strains included 14 belonged to ST8, 10 to ST1, 5 to ST5, and one each to ST15, ST20, ST97, ST291, ST1516, ST2725, ST2764, and ST4143 (Table S1).

### Sequencing by MiSeq and Analysis

DNA was extracted from *S. aureus* using achromopeptidase and Phenol/Chloroform/Isoamyl Alcohol (25:24:1)(FUJIFILM Wako Pure Chemical Corporation, Osaka, Japan) and purified using the spin column method with the FastGene Gel/PCR Extraction Kit (NIPPON Genetics Co, Ltd, Tokyo, Japan). DNA library preparation was performed using the Illumina DNA Prep (M) Tagmentation Library Preparation kit and IDT for Illumina DNA/RNA UD Indexes (Illumina). Sequencing was planned with 300 bp paired-end reads using the 600-cycle MiSeq Reagent Kit v3 (Illumina). Reads were trimmed and filtered using Trimmomatic version 0.39^30^, applying a quality score of 20 and a minimum read length of 50 bp, and de novo assembly to generate contigs was performed using SPAdes version 3.15.1.^31^

### Sequencing by MinION R10.4.1 and Analysis

High molecular weight DNA was extracted from *S. aureus* strains by bead-beating with Ez-Beas (Promega K.K., Tokyo, Japan) and processed with the magLEAD system and the magDEA SV and PS protocol (Precision System Science Co., Ltd., Chiba, Japan). For ONT, DNA library preparation was performed using the Rapid Barcoding Kit 24 V14 SQK-RBK114.24 (ONT). Sequencing was conducted on MinION with Flow Cell (R10.4.1) FLO-MIN114 (ONT) for up to 72 hours. Basecalling and demultiplexing were performed using Dorado version 0.6.0 with the dna_r10.4.1_e8.2_400bps_sup@v4.1.0 model. Reads were trimmed and filtered with NanoFilt version 2.8.0, applying a quality score of 10 and a minimum read length of 1,000 bp.^32^ To limit downstream data volume, reads were downsampled with Seqkit version 2.6.1, ensuring that the total nucleotide count did not exceed 200 times the genome size of *S. aureus*.^33^ Self-error correction was carried out using DeChat version 1.0.1, and de novo assembly was performed with Flye version 2.9.4.^34,35^

### Species Identification, Basic Typing, and Gene Identification

Species identification was primarily conducted using KmerFinder, and subsequently confirmed by calculating the Average Nucleotide Identity against the type strain genome using FastANI.^36,37^ A threshold of 95.0% ANI was applied as the cutoff for species identification.^38^ Identification of acquired AMR genes and nucleotide point mutations in the quinolone resistance-determining regions (QRDRs) of DNA gyrase subunit A (GyrA) and DNA topoisomerase IV subunit A (GrlA), as well as the rifampicin resistance-determining regions (RRDRs) of RNA polymerases subunit B (RpoB), was conducted with ResFinder version 4.5.0.^39^ SCC*mec* typing was performed using SCCmecFinder version 1.0.0.^40^ MLST was conducted following the PubMLST scheme (https://pubmlst.org/organisms/staphylococcus-aureus). Virulence gene identification was performed with VirulenceFinder version 2.0.5.^41^ These analyses were conducted on genomes generated from both Illumina and ONT sequencing. Default parameters were used for all tools in this section.

### Successive Core-genome SNP-based Phylogenetic Analysis in TAROT

TAROT was validated by randomly inputting MinION data one at a time into the workflow and comparing it to Illumina data inputs. The TAROT workflow is illustrated in Figure 2. Representative STs of MRSA reference genomes were selected based on a review article and prepared within the TAROT database (Table S2).^42^ For mapping analysis, a reference genome was selected based on either MLST analysis results using stringMLST^43^ or the best mapping outcome across multiple references using BBsplit (version was last modified June 11, 2018), a tool included in BBTools (https://github.com/kbaseapps/BBTools). Simulations of short reads for 300 bp of 1,000,000 paired-end reads were generated using Wgsim version 1.20.^44^ Mapping to the reference genome and generating BAM files was performed with BWA version 0.7.17 using the sw option, and SAMtools version 1.20.^44,45^ The BAM files were grouped and organized based on MLST or BBsplit results. The core-genome was extracted from the grouped BAM files using SAMtools with the mpileup option and VarScan version 2.3.9 with the mpileup2cns option.^46^ Recombination regions in the core-genome were identified and excluded using ClonalFrameML version 1.12, followed by text editing in the Bash console.^47^ Pairwise cgSNP calculations were performed using SNP-dists version 0.8.2 (https://github.com/tseemann/snp-dists). Phylogenetic trees were generated using RAxML version 8.2.12 with the raxmlHPC-PTHREADS option, model GAMMAMI, and 1000 bootstrap replications.^48^ The entire time for inputting FASTQ files into TAROT and outputting a phylogenetic tree was measured using the time command on Ubuntu 20.04.

**Figure 2.**
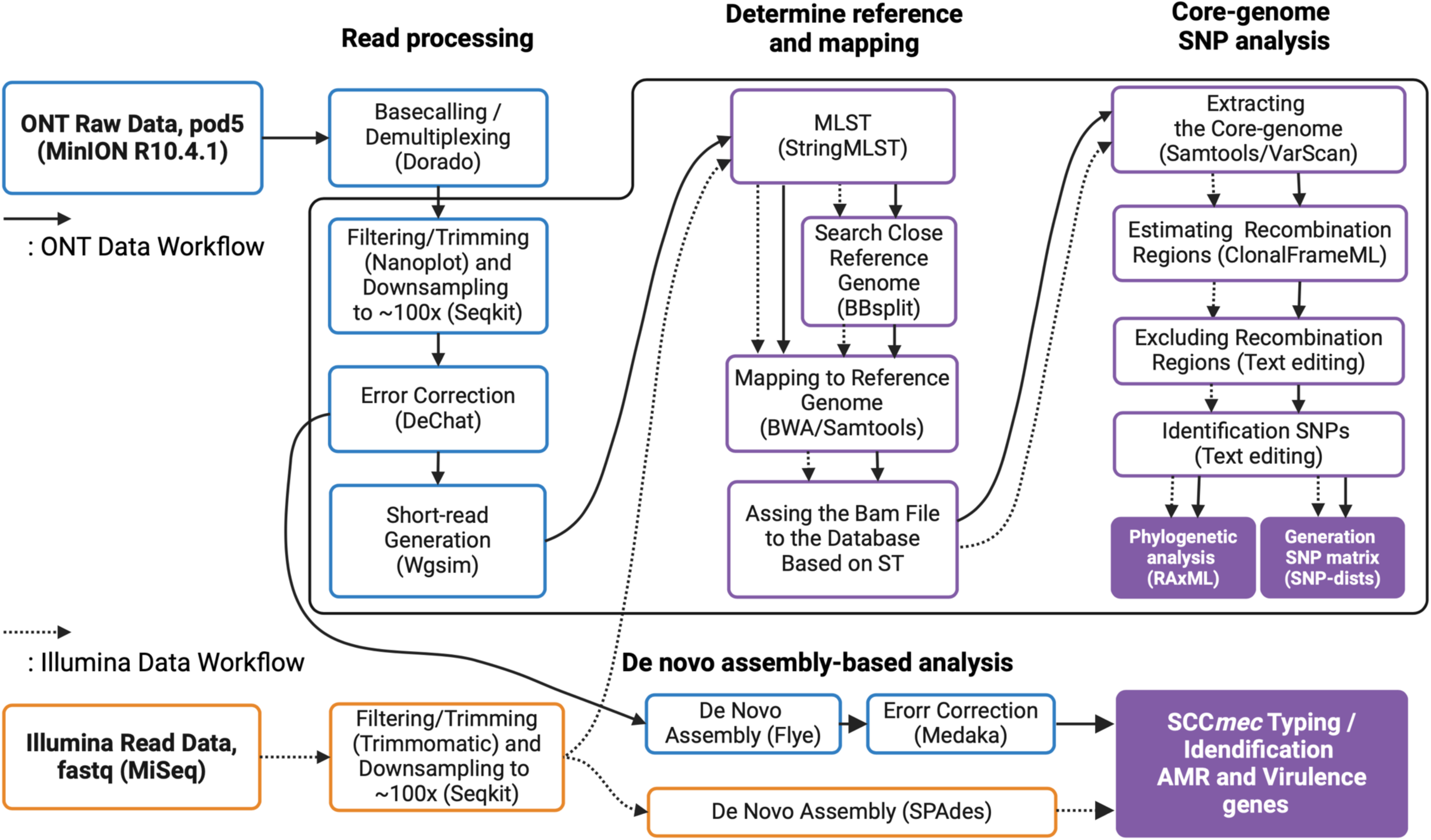
The Bioinformatics Workflow of Tracking Antimicrobial Resistance Organisms Timely (TAROT). The blue-outlined box indicates the processes specific to Oxford Nanopore Technologies (ONT). The orange-outlined box shows processes unique to Illumina data. The purple-outlined box shows processes for both ONT and Illumina data, handled separately. The purple box marks the destination of the workflow. Solid arrows represent the ONT data workflow, while dashed arrows indicate the Illumina data workflow. The time taken for the workflow in the solid box processes was measured using the ‘time’ command in Linux.

### Data availability

The WGS data have been deposited in GenBank under the BioProject accession number PRJNA1090044, and SRA and nucleotide accession numbers are shown in Table S1.

### Code Availability

The TAROT code is available on GitHub (https://github.com/KotaroAoki/TAROT_MRSA) and can be run in a Docker container (https://hub.docker.com/repository/docker/kotaroaoki/tarot-mrsa-env/general).

## Results

### Sequencing Yield and Assembly Results of MinION and MiSeq

The average sequencing depth per MinION run for the ST8 reference genome, obtained from sequencing 34 strains across four runs, was 73x at 1 hour, 262x at 3 hours, 451x at 6 hours, 1,901x at 24 hours, and 3,080x at 48 hours (Table S3). The average sequencing depths achieved by MinION and MiSeq for each assembled genome were 65x and 98x, respectively (Table S1). Assembly using ONT reads provided closed genome replicons in 30 of the 34 strains. Among these, two closed plasmids were detected in six strains, and one closed plasmid in 18 strains (Table S1). The results of MLST and SCC*mec* typing showed strict concordance between MinION and MiSeq (Table S1).

### Successive phylogenetic Analysis Using TAROT-ONT

A successive phylogenetic analysis using TAROT-ONT generated 11 trees for 14 strains of ST8, seven trees for ten strains of ST1, and two trees for five strains of ST5 (Figure 3A to 3C). TAROT-ONT assigned MRSA strains to the same ST as those identified by Illumina. Here, we set the threshold for highly suspected transmission as pairwise cgSNP < 5 and for suspected transmission as pairwise cgSNP < 15. Four pairs of highly suspected transmission events were detected in ST8 (Figure 3A and Table S4), four in ST1 (Figure 3B and Table S4), and two in ST5 (Figure 3C and Table S4). In ST8, three suspected transmission events were identified, with TUM22721 added in tree #11 related to one of the highly suspected transmission pairs. Timely detection of highly suspected or suspected transmissions occurred in trees #6 through #11 for ST8, trees #3, #5, and #7 for ST1, and tree #2 for ST5.

**Figure 3.**
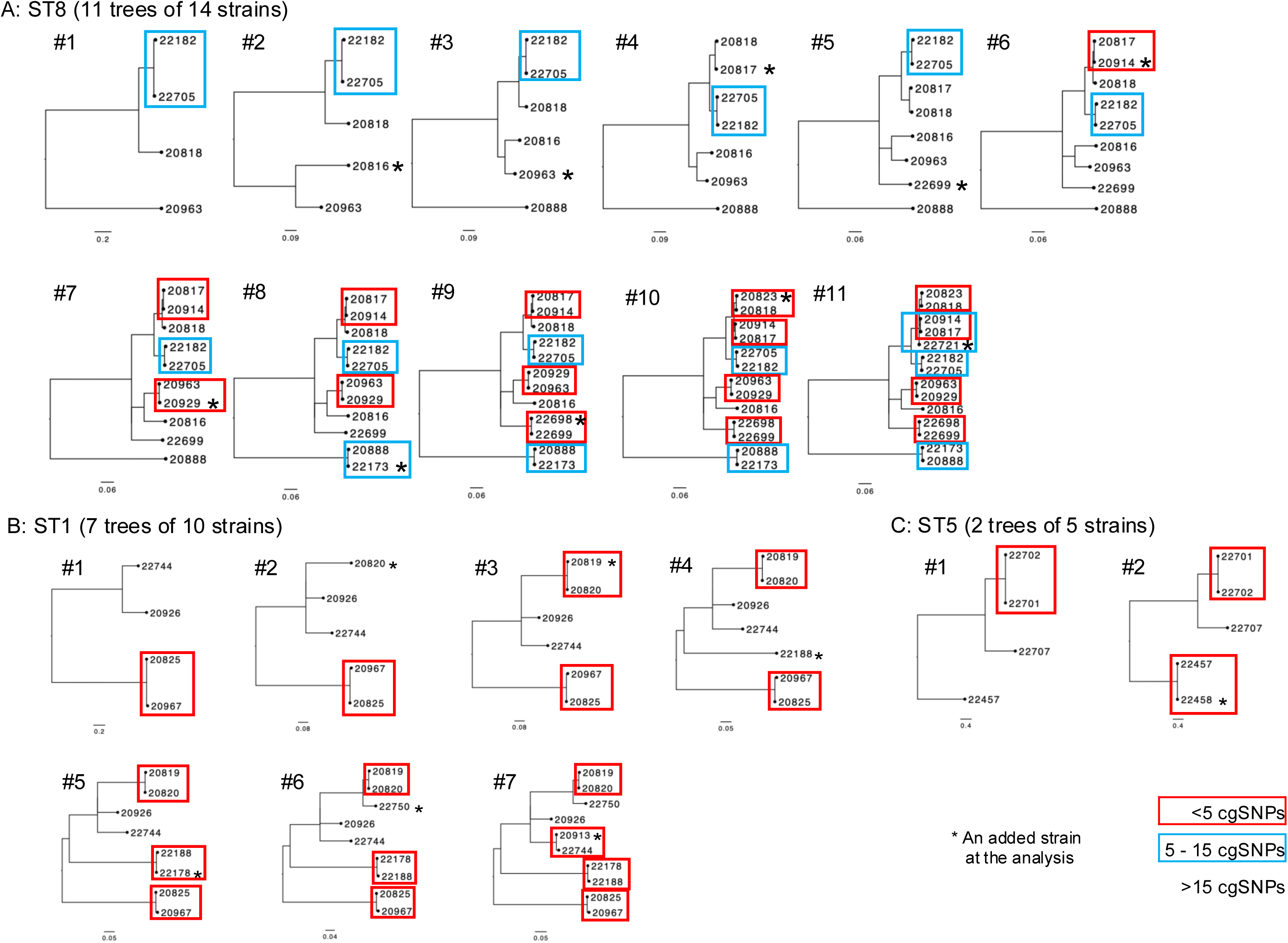
Phylogenetic Trees Obtained by Successive Core-Genome SNP (cgSNP)-Based Phylogenetic Analysis by TAROT-ONT. A, Phylogenetic trees for ST8; B, Phylogenetic trees for ST1; C, Phylogenetic trees for ST1. The red box highlights transmission pairs highly suspected to have fewer than five cgSNPs. The blue box highlights transmission pairs suspected to range from five to 15 cgSNPs. Each asterisk denotes a strain added to the analysis.

Apart from strains belonging to ST8, ST1, and ST5, TUM22730 belonged to ST97 and was assigned to a reference genome of the same ST in the database (Table S5). The remaining four strains were not assigned to any reference genome in the database by MLST and were classified using BBsplit as a reference genome classifier. TUM20915 belonged to ST2725, TUM22749 to ST2764, and TUM22741 to ST4143, which were all classified as ST1_bbsplit. ST1_bbsplit differed from ST1 and was obviously unrelated to nosocomial transmissions; therefore, these strains were not included in cgSNP-based phylogenic analysis of ST1 strains. TUM22723, belonging to ST1516, was assigned to ST228_bbsplit.

### Comparison of Core-Genome SNP-based Phylogenetic Analysis Accuracy Between TAROT-ONT and Illumina

The proportion of core-genome size over the reference genome, calculated using the maximum number of strains in cgSNP-based phylogenetic analysis for TAROT-ONT and Illumina, were 93.1% and 94.3% for ST8, 97.2% and 98.6% for ST1, and 95.3% and 97.6% for ST5, respectively (Table S5). The differences in cgSNPs identified between TAROT-ONT and Illumina ranged from zero to 17 for ST8, zero to 14 for ST1, and zero to 5 for ST5 (Figure 4). In pairs of cgSNPs with fewer than 100 identified in Illumina, the differences were significantly lower, ranging from zero to two for ST8, zero to eight for ST1, and zero to two for ST5. Additionally, in pairs of cgSNPs with fewer than 20 in Illumina, the differences ranged from zero to two for ST8 and were zero for ST1 and ST5.

**Figure 4.**
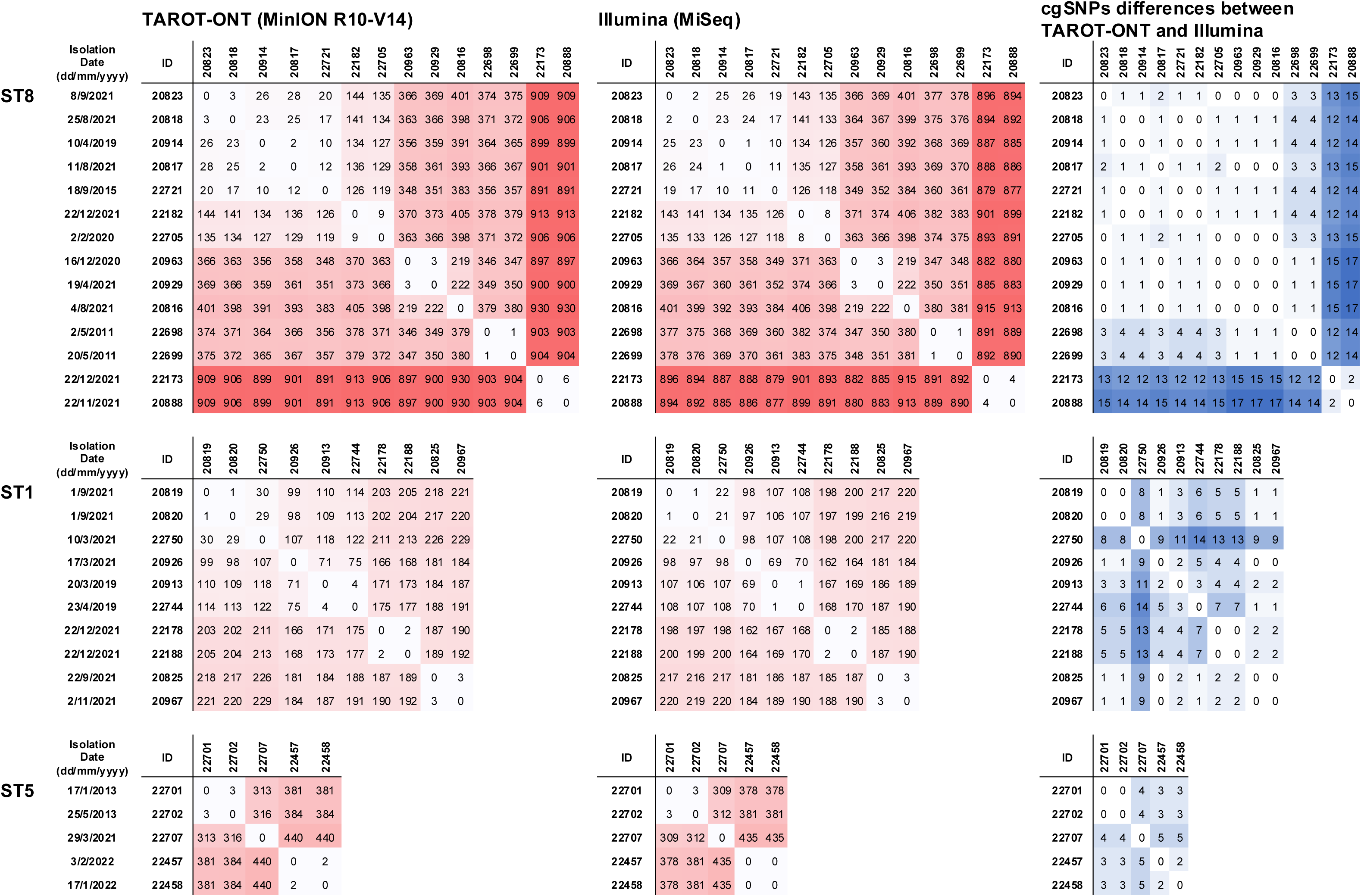
Comparison of Core-genome SNP (cgSNP) Identification Differences Between TAROT-ONT and Illumina. The left and middle columns display matrices showing pairwise cgSNP values, while the right column illustrates the differences in cgSNP values between TAROT-ONT and Illumina. Darker colours in the heatmap indicate higher cgSNP values.

### Time Taken for Each Bioinformatic Analysis of Each Strain in TAROT-ONT

The time required for the bioinformatic analysis of each strain, from inputting FASTQ data from ONT to obtaining a phylogenetic tree via the TAROT workflow, ranged from five to 42 minutes (Table S5). Analysis times were significantly shorter when three or fewer strains were assigned to the same reference genome, as phylogenetic analysis was not conducted in those cases. The time required increased with the number of strains assigned to the same reference genome.

### Comparison of Detection for Genes of Interest Between ONT and Illumina

Using ResFinder with contigs obtained from ONT or Illumina, ten types of acquired AMR genes were identified in 34 strains (Figure 5). The only false-negative result for ONT, using Illumina as the reference, was *blaZ* in TUM22741. In TUM22741, *blaZ* was detected as part of a gene sequence comprising 681 bp out of the total gene length of 864 bp. The average ONT sequence depth for the contigs of TUM22741 was 19x, which was slightly low (Table S1). Conversely, the false-positive results detected by ONT included *blaZ* in TUM22701, TUM22702, TUM22457, and TUM22458, as well as *ermA* in TUM22701 and TUM22702, all of which were detected only as partial gene length by Illumina (Figure 5). ONT detected multiple copies of resistance genes such as *aac(6’)-aph(2’’)*, *ant(9)-Ia*, *blaZ, cat,* and *ermA* in several strains (Figure 5). There was no discrepancy between ONT and Illumina in identifying QRDR mutations in GyrA and GrlA, RRDR mutations in RpoB, and virulence genes (Figures 5 and 6).

**Figure 5.**
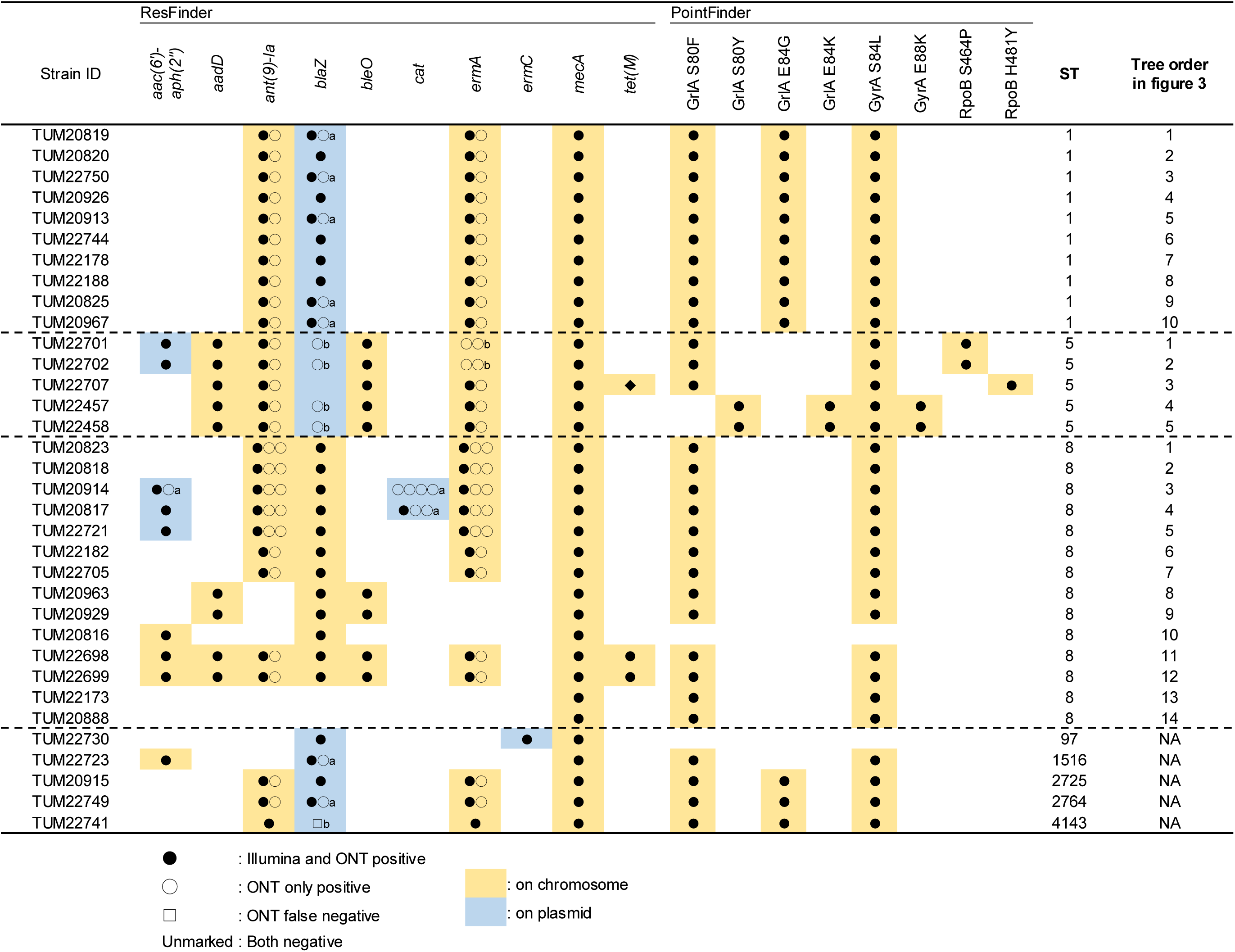
Comparison of Antimicrobial Resistance Genes and Mutation Identification Between ONT and Illumina. Antimicrobial resistance genes and mutations were identified using ResFinder. The number of symbols indicates the gene copy number in each strain. a, Assembly errors probably resulted in false multi-copy gene detection; b, Partial gene sequence detected. In TUM22741, a partial *blaZ* sequence was detected by ONT data. In TUM22701 and TUM22702, partial *blaZ* and *ermA* sequences were detected by Illumina. In TUM22457 and TUM22458, Illumina detected partial *ermA* sequences. ST stands for sequence type of multilocus sequence typing; E, Glutamic Acid; F, Phenylalanine; G, Glycine; H, Histidine; K, Lysine; L, Leucine; P, Proline; S, Serine; Y, Tyrosine.

**Figure 6.**
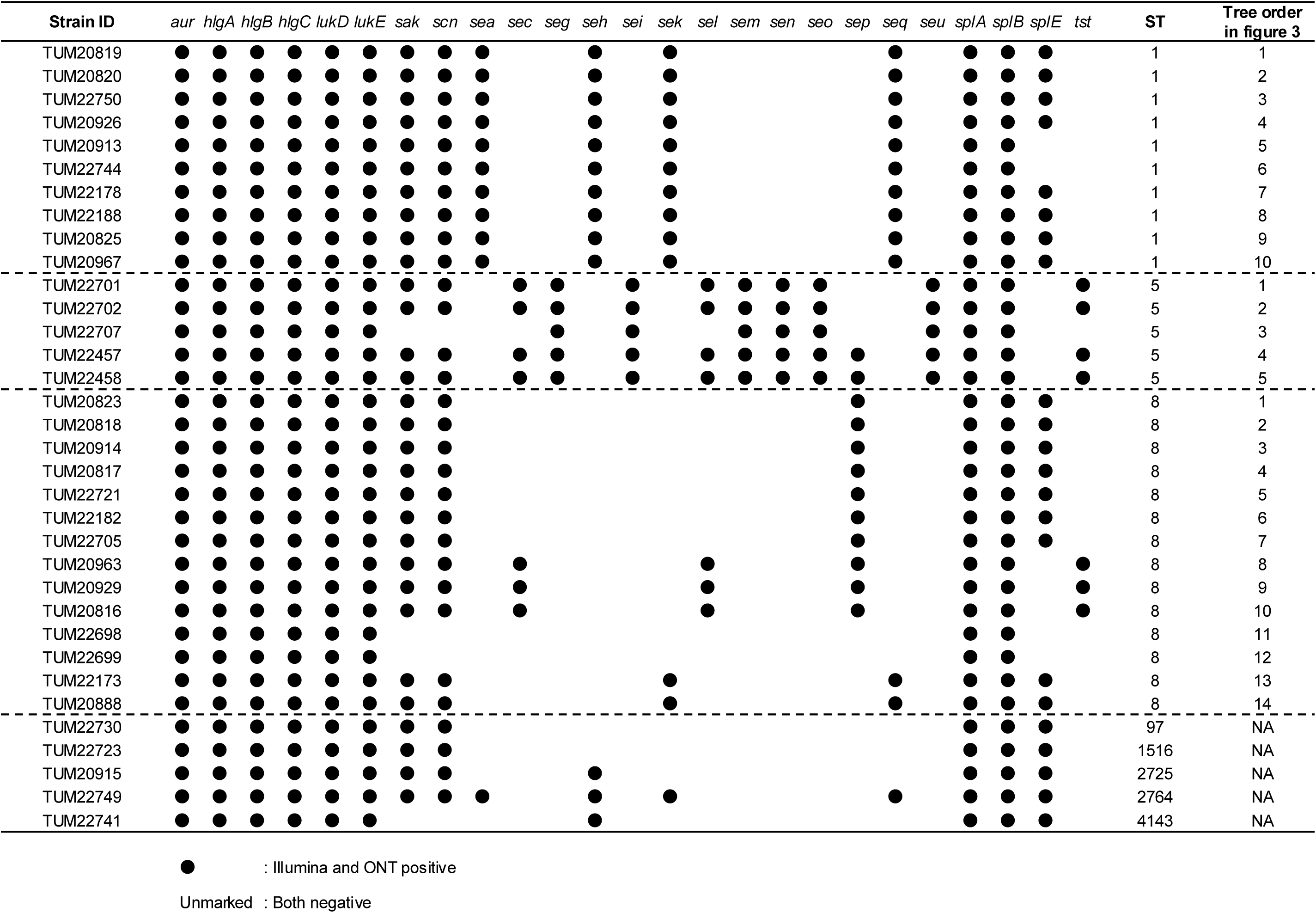
Comparison of Virulence Genes Between ONT and Illumina. Virulence genes were identified using VirulenceFinder. ST stands for sequence type of multilocus sequence typing.

### Comparison of Genome Structure for Single or Multi-copy AMR Gene-Detected Replicons

A part of multi-copy gene detection probably results in assembly errors. Two types of *blaZ*-carrying circular plasmids were identified in ST1 strains (Figure 5). One type of plasmid, carrying a single *blaZ* (approximately 21,326 bp), was identified in five strains (TUM20926, TUM20820, TUM22178, TUM22744, and TUM22188)(Figure S1). The other type of plasmid, carrying two copies of *blaZ* (approximately 42,652 bp), was identified in five additional strains (TUM20913, TUM20819, TUM22750, TUM20967, and TUM20825)(Figure S1). The plasmid carrying two copies of the *blaZ* consisted of two single-copy *blaZ*-carrying plasmids connected in tandem. The smaller plasmids carrying a single *blaZ* appeared to be a structures unit (Figure S1). Mapping analyses to confirm whether assembly errors of ONT data generated the tandem unit sequences revealed no reads longer than the unit structure length (approximately 21,326 bp) in plasmid-harboring strains with two copies of *blaZ*. In 10 ST8 strains, *blaZ* was located on the chromosome as a single copy (Figure 5).

A plasmid carrying two copies of *aac(6’)-aph(2’’)* in strain TUM20914 (pMTY20914-1) was composed of a tandemly connected plasmid carrying a single copy of *aac(6’)-aph(2’’)* in TUM20817 (pMTY20817-1)(Figure 5 and S2). Mapping analysis of ONT reads from TUM20914 to pMTY20914-1 found only shorter reads than the length of pMTY20817-1. Another plasmid type, carrying multiple copies of *cat* in TUM20817 (pMTY20817-2) and TUM20914 (pMTY20914-2), consisted of a single unit of approximately 4.2 kb, with three and four copies connected in tandem, respectively (Figure 5 and S3). Like blaZ-carrying plasmids, mapping analysis of ONT reads from TUM20817 and TUM20914 to pMTY20817-2 and pMTY20914-2 found only shorter reads than the length of the single unit.

*ermA* and *ant(9)-Ia* were located adjacent to each other on the chromosomes of 27 strains. This gene pair was a single copy in one strain, two copies in 21 strains, and three copies in five strains (Figure 5). These gene pairs were located within an approximately 6.3 kb unit containing integrase genes. In comparative chromosome structure analysis of representative strains TUM22741 (single-copy), TUM22182 (two-copy), and TUM22721 (three-copy), the insertion positions of the first and second units were identical among the strains (Figure S4).

## Discussions

TAROT has been developed to provide timely feedback on nosocomial transmission of AMR organisms for IPC intervention, using ONT technology instead of batch WGS bacteria processing via Illumina in central laboratories. Recent updates have enhanced ONT performance to a level comparable to Illumina, making the TAROT concept feasible. ONT sequencers offer strong scalability and an optimal balance between consumable costs and throughput. For instance, MinION provides adequate throughput to analyze approximately 12 MRSA strains in a single run. If only one strain is analyzed, the Flongle can be used instead to save consumable costs. Regarding data analysis, clinical microbiology laboratories in hospitals generally employ few bioinformaticians, so bioinformatics support is essential. Automating the data analysis workflow helps reduce the workload on clinical laboratory staff, enables timely feedback to IPC teams, and contributes to standardization and quality control of the entire process.

The advantage of the TAROT concept over batch WGS and cgSNP-based phylogenetic analysis is its ability to analyze each strain with ONT and cgSNP methods at a quality level comparable to Illumina, providing feedback within the same day.^25^ In a previous study, batch WGS of MRSA and cgSNP-based analysis identified an outbreak within 33 hours starting from strain processing.^49^ However, batch processing in IPC intervention requires waiting to accumulate sufficient samples for a batch, delaying response time. Validated in this study, TAROT provides a faster turnaround time (TAT), enabling timely feedback that can efficiently inform IPC intervention at the bedside. The entire TAROT process, from DNA extraction of MRSA to the phylogenetic analysis report, can be completed within six hours. The breakdown is as follows: one hour for DNA extraction, one hour for ONT library preparation, three hours for ONT sequencing, and one hour for bioinformatic analysis. The average time to achieve a 50x sequence depth per MRSA genome was 44 minutes (Table S3), potentially allowing the TAROT’s TAT to be less than six hours. If Flongle, with lower throughput, is used instead of MinION, approximately three hours should be allocated for sequencing.

A prospective genomic surveillance study of MRSA in NICU patients using batch sample processing with Illumina HTS demonstrated its utility by identified local transmission events.^50^ Genome analyses using ONT is valuable for outbreak investigations not only for MRSA, as used in this study but also for Gram-negative bacteria such as Shiga toxin-producing *Escherichia coli*, described as a tool for ‘closing the gap’ from the short-read sequencing era.^25^ The updated ONT R10.4.1, which was also used in this study and offers higher nucleotide sequencing accuracy compared to R9.4.1, has proven valuable for bacterial genomic epidemiology.^26,51,52^ Our ONT results showed that AMR and virulence gene identification, as well as MLST and SCC*mec* typing results, were highly concordant with Illumina results. However, contigs identified as plasmids carrying AMR genes in certain strains suggested possible assembly errors with the Flye assembler. Comparative plasmid structure analyses using ONT-only de novo assemblies may still require visual inspection of read mapping to the generated contigs. Notably, the assembly process branched from the TAROT workflow to minimize the time required for high-priority phylogenetic analysis (Figure 2).

An appropriate threshold of cgSNPs to rule out nosocomial transmission of clonal isolates could support effective IPC interventions. Coll et al. proposed a conservative cgSNP threshold of 15 to exclude transmission within six months, considering within-host strain variation and evolutionary rate, based on WGS data from MRSA strains isolated from 455 patients using Illumina.^17^ According to this threshold, 95% of epidemiologically linked cases could be identified among 775 cases.^17^ TAROT-ONT showed high compatibility with Illumina when pairwise cgSNP values were <20, suggesting that this threshold could be applied to TAROT-ONT interpretations. Successive MRSA analyses using TAROT-ONT in high risk wards for nosocomial transmission can timely identify MRSA strains that require specific transmission prevention interventions. TAROT could help reduce staff workload by enabling focused IPC interventions. This study has four potential limitations. First, the number of strains and the selection method used to validate TAROT were limited. TAROT could be further enhanced to provide a more robust workflow by analyzing more strains, especially when applied for MRSA IPC interventions in hospitals. Second, TAROT’s phylogenetic analysis results were not evaluated based on epidemiological links in a hospital setting. TAROT could prioritize cases by identifying MRSA strains with a higher transmission likelihood based on molecular epidemiology. Therefore, future studies should consider evaluating this functionality. Third, we simulated successive phylogenetic analyses using in silico methods that accounted for ONT sequencing consumable kits. Users can choose MinION or Flongle depending on the number of strains analyzed daily. Fourth, while we have published a TAROT script and a Docker container image to build the necessary environment, we have not yet developed web services for TAROT. Such services would be essential for deploying TAROT in hospitals, and we plan to develop a network architecture for authorized members to have secure access.

In conclusion, TAROT-ONT provides accuracy compatible with cgSNP-based phylogenetic analysis using Illumina for MRSA, facilitating effective IPC interventions in hospitals by offering timely feedback.

## Data Availability

https://github.com/KotaroAoki/TAROT_MRSA

https://hub.docker.com/repository/docker/kotaroaoki/tarot-mrsa-env/general

## Acknowledgments

We thank Dr. Yoshikazu Ishii for advising us about conducting this study.

## Transparency declarations

SH reports speaker payments from MSD, Shionogi, Pfizer and consulting fees from Shionogi, Pfizer, Denka outside the submitted work. All other authors have no conflicts of interest to declare.

## Ethics declaration

We used MRSA strains isolated from patients, but we did not use any information to identify individuals. This study has been approved by the Thesis Compliance Committee of Toho University School of Medicine as having been conducted through appropriate legal procedures.

## References

1. Zhen X, Lundborg CS, Zhang M, et al. Clinical and economic impact of methicillin-resistant Staphylococcus aureus: a multicentre study in China. Sci Rep 2020; 10: 3900.

2. Zhen X, Stålsby Lundborg C, Sun X, Zhu N, Gu S, Dong H. Economic burden of antibiotic resistance in China: a national level estimate for inpatients. Antimicrob Resist Infect Control 2021; 10: 5.

3. Zhen X, Stålsby Lundborg C, Sun X, Hu X, Dong H. Clinical and economic impact of third-generation cephalosporin-resistant infection or colonization caused by Escherichia coli and Klebsiella pneumoniae: A multicenter study in China. Int J Environ Res Public Health 2020; 17: 9285.

4. Rocha LEC, Singh V, Esch M, Lenaerts T, Liljeros F, Thorson A. Dynamic contact networks of patients and MRSA spread in hospitals. Sci Rep 2020; 10: 9336.

5. Pei S, Liljeros F, Shaman J. Identifying asymptomatic spreaders of antimicrobial-resistant pathogens in hospital settings. Proc Natl Acad Sci U S A 2021; 118: e2111190118.

6. Ogihara S, Yamaguchi T, Sato T, et al. Assessing the discriminability of PCR-based open reading frame typing versus single-nucleotide polymorphism analysis via draft whole-genome sequencing of methicillin-resistant Staphylococcus aureus in nosocomial transmission analysis. J Infect Chemother 2024; 30: 951–4.

7. Neoh H-M, Tan X-E, Sapri HF, Tan TL. Pulsed-field gel electrophoresis (PFGE): A review of the ‘gold standard’ for bacteria typing and current alternatives. Infect Genet Evol 2019; 74: 103935.

8. Kao CM, Fritz SA. Infection prevention-how can we prevent transmission of community-onset methicillin-resistant Staphylococcus aureus? Clin Microbiol Infect 2024. Available at: 10.1016/j.cmi.2024.01.004.

9. Uelze L, Grützke J, Borowiak M, et al. Typing methods based on whole genome sequencing data. One Health Outlook 2020; 2: 3.

10. Popovich KJ, Snitkin ES. Whole genome sequencing-implications for infection prevention and outbreak investigations. Curr Infect Dis Rep 2017; 19: 15.

11. Popovich KJ, Aureden K, Ham DC, et al. SHEA/IDSA/APIC Practice Recommendation: Strategies to prevent methicillin-resistant Staphylococcus aureus transmission and infection in acute-care hospitals: 2022 Update. Infect Control Hosp Epidemiol 2023; 44: 1039–67.

12. Popovich KJ, Green SJ, Okamoto K, et al. MRSA transmission in intensive care units: Genomic analysis of patients, their environments, and healthcare workers. Clin Infect Dis 2021; 72: 1879–87.

13. Cheng VCC, Wong S-C, Cao H, et al. Whole-genome sequencing data-based modeling for the investigation of an outbreak of community-associated methicillin-resistant Staphylococcus aureus in a neonatal intensive care unit in Hong Kong. Eur J Clin Microbiol Infect Dis 2019; 38: 563–73.

14. Bartels MD, Petersen A, Worning P, et al. Comparing whole-genome sequencing with Sanger sequencing for spa typing of methicillin-resistant Staphylococcus aureus. J Clin Microbiol 2014; 52: 4305–8.

15. Stevens EL, Carleton HA, Beal J, et al. Use of whole genome sequencing by the federal Interagency Collaboration for Genomics for food and Feed Safety in the United States. J Food Prot 2022; 85: 755–72.

16. Köser CU, Holden MTG, Ellington MJ, et al. Rapid whole-genome sequencing for investigation of a neonatal MRSA outbreak. N Engl J Med 2012; 366: 2267–75.

17. Coll F, Raven KE, Knight GM, et al. Definition of a genetic relatedness cutoff to exclude recent transmission of meticillin-resistant Staphylococcus aureus: a genomic epidemiology analysis. Lancet Microbe 2020; 1: e328–35.

18. Blane B, Raven KE, Brown NM, et al. Evaluating the impact of genomic epidemiology of methicillin-resistant Staphylococcus aureus (MRSA) on hospital infection prevention and control decisions. Microb Genom 2024; 10. Available at: https://pubmed.ncbi.nlm.nih.gov/38630616/.

19. Kubota KA, Wolfgang WJ, Baker DJ, et al. PulseNet and the changing paradigm of laboratory-based surveillance for foodborne diseases. Public Health Rep 2019; 134: 22S–28S.

20. Michelacci V, Asséré A, Cacciò S, et al. European Union Reference Laboratories support the National food, feed and veterinary Reference Laboratories with rolling out whole genome sequencing in Europe. Microb Genom 2023; 9. Available at: 10.1099/mgen.0.001074.

21. Blane B, Raven KE, Leek D, Brown N, Parkhill J, Peacock SJ. Rapid sequencing of MRSA direct from clinical plates in a routine microbiology laboratory. J Antimicrob Chemother 2019; 74: 2153–6.

22. Madera S, McNeil N, Serpa PH, et al. Prolonged silent carriage, genomic virulence potential and transmission between staff and patients characterize a neonatal intensive care unit (NICU) outbreak of methicillin-resistant Staphylococcus aureus (MRSA). Infect Control Hosp Epidemiol 2023; 44: 40–6.

23. Reuter S, Ellington MJ, Cartwright EJP, et al. Rapid bacterial whole-genome sequencing to enhance diagnostic and public health microbiology. JAMA Intern Med 2013; 173: 1397–404.

24. Wang Y, Zhao Y, Bollas A, Wang Y, Au KF. Nanopore sequencing technology, bioinformatics and applications. Nat Biotechnol 2021; 39: 1348–65.

25. Bogaerts B, Van den Bossche A, Verhaegen B, et al. Closing the gap: Oxford Nanopore Technologies R10 sequencing allows comparable results to Illumina sequencing for SNP-based outbreak investigation of bacterial pathogens. J Clin Microbiol 2024; 62: e0157623.

26. Mostafa HH. An evolution of Nanopore next-generation sequencing technology: implications for medical microbiology and public health. J Clin Microbiol 2024; 62: e0024624.

27. Ni Y, Liu X, Simeneh ZM, Yang M, Li R. Benchmarking of Nanopore R10.4 and R9.4.1 flow cells in single-cell whole-genome amplification and whole-genome shotgun sequencing. Comput Struct Biotechnol J 2023; 21: 2352–64.

28. Raven KE, Bragin E, Blane B, et al. Large-scale evaluation of a rapid fully automated analysis platform to detect and refute outbreaks based on MRSA genome comparisons. mSphere 2022; 7: e0028322.

29. Brown NM, Blane B, Raven KE, et al. Pilot evaluation of a fully automated bioinformatics system for analysis of methicillin-resistant Staphylococcus aureus genomes and detection of outbreaks. J Clin Microbiol 2019; 57. Available at: 10.1128/JCM.00858-19.

30. Bolger AM, Lohse M, Usadel B. Trimmomatic: a flexible trimmer for Illumina sequence data. Bioinformatics 2014; 30: 2114–20.

31. Bankevich A, Nurk S, Antipov D, et al. SPAdes: a new genome assembly algorithm and its applications to single-cell sequencing. J Comput Biol 2012; 19: 455–77.

32. De Coster W, D’Hert S, Schultz DT, Cruts M, Van Broeckhoven C. NanoPack: visualizing and processing long-read sequencing data. Bioinformatics 2018; 34: 2666–9.

33. Shen W, Le S, Li Y, Hu F. SeqKit: A cross-platform and ultrafast toolkit for FASTA/Q file manipulation. PLoS One 2016; 11: e0163962.

34. Li Y, Chen E, Xu J, et al. Repeat and haplotype aware error correction in nanopore sequencing reads with DeChat. bioRxiv 2024: 2024.05.09.593079. Available at: https://www.biorxiv.org/content/biorxiv/early/2024/05/10/2024.05.09.593079. Accessed May 13, 2024.

35. Kolmogorov M, Yuan J, Lin Y, Pevzner PA. Assembly of long, error-prone reads using repeat graphs. Nat Biotechnol 2019; 37: 540–6.

36. Larsen MV, Cosentino S, Lukjancenko O, et al. Benchmarking of methods for genomic taxonomy. J Clin Microbiol 2014; 52: 1529–39.

37. Jain C, Rodriguez-R LM, Phillippy AM, Konstantinidis KT, Aluru S. High throughput ANI analysis of 90K prokaryotic genomes reveals clear species boundaries. Nat Commun 2018; 9: 5114.

38. Richter M, Rosselló-Móra R. Shifting the genomic gold standard for the prokaryotic species definition. Proc Natl Acad Sci U S A 2009; 106: 19126–31.

39. Bortolaia V, Kaas RS, Ruppe E, et al. ResFinder 4.0 for predictions of phenotypes from genotypes. J Antimicrob Chemother 2020; 75: 3491–500.

40. Kaya H, Hasman H, Larsen J, et al. SCCmecFinder, a Web-Based Tool for Typing of Staphylococcal Cassette Chromosome mec in Staphylococcus aureus Using Whole-Genome Sequence Data. mSphere 2018; 3. Available at: 10.1128/mSphere.00612-17.

41. Malberg Tetzschner AM, Johnson JR, Johnston BD, Lund O, Scheutz F. In silico genotyping of Escherichia coli isolates for extraintestinal virulence genes by use of whole-genome sequencing data. J Clin Microbiol 2020; 58. Available at: 10.1128/JCM.01269-20.

42. Lakhundi S, Zhang K. Methicillin-Resistant Staphylococcus aureus: Molecular Characterization, Evolution, and Epidemiology. Clin Microbiol Rev 2018; 31. Available at: 10.1128/CMR.00020-18.

43. Gupta A, Jordan IK, Rishishwar L. stringMLST: a fast k-mer based tool for multilocus sequence typing. Bioinformatics 2017; 33: 119–21.

44. Li H, Handsaker B, Wysoker A, et al. The Sequence Alignment/Map format and SAMtools. Bioinformatics 2009; 25: 2078–9.

45. Li H, Durbin R. Fast and accurate short read alignment with Burrows-Wheeler transform. Bioinformatics 2009; 25: 1754–60.

46. Koboldt DC, Zhang Q, Larson DE, et al. VarScan 2: somatic mutation and copy number alteration discovery in cancer by exome sequencing. Genome Res 2012; 22: 568–76.

47. Didelot X, Wilson DJ. ClonalFrameML: efficient inference of recombination in whole bacterial genomes. PLoS Comput Biol 2015; 11: e1004041.

48. Stamatakis A. RAxML version 8: a tool for phylogenetic analysis and post-analysis of large phylogenies. Bioinformatics 2014; 30: 1312–3.

49. Ferreira FA, Helmersen K, Visnovska T, Jørgensen SB, Aamot HV. Rapid nanopore-based DNA sequencing protocol of antibiotic-resistant bacteria for use in surveillance and outbreak investigation. Microb Genom 2021; 7. Available at: 10.1099/mgen.0.000557.

50. Worley JN, Crothers JW, Wolfgang WJ, et al. Prospective genomic surveillance reveals cryptic MRSA outbreaks with local to international origins among NICU patients. J Clin Microbiol 2023; 61: e0001423.

51. Sanderson ND, Hopkins KMV, Colpus M, et al. Evaluation of the accuracy of bacterial genome reconstruction with Oxford Nanopore R10.4.1 long-read-only sequencing. Microb Genom 2024; 10. Available at: 10.1099/mgen.0.001246.

52. Ye L, Liu X, Ni Y, et al. Comprehensive genomic and plasmid characterization of multidrug-resistant bacterial strains by R10.4.1 nanopore sequencing. Microbiol Res 2024; 283: 127666.

