## Supplementary figures and images for "Tracking Antimicrobial Resistant Organisms Timely (TAROT): A Workflow Validation Study for Successive Core-genome SNP-based Nosocomial Transmission Analysis"

### Supplemental Figure 1 to 4

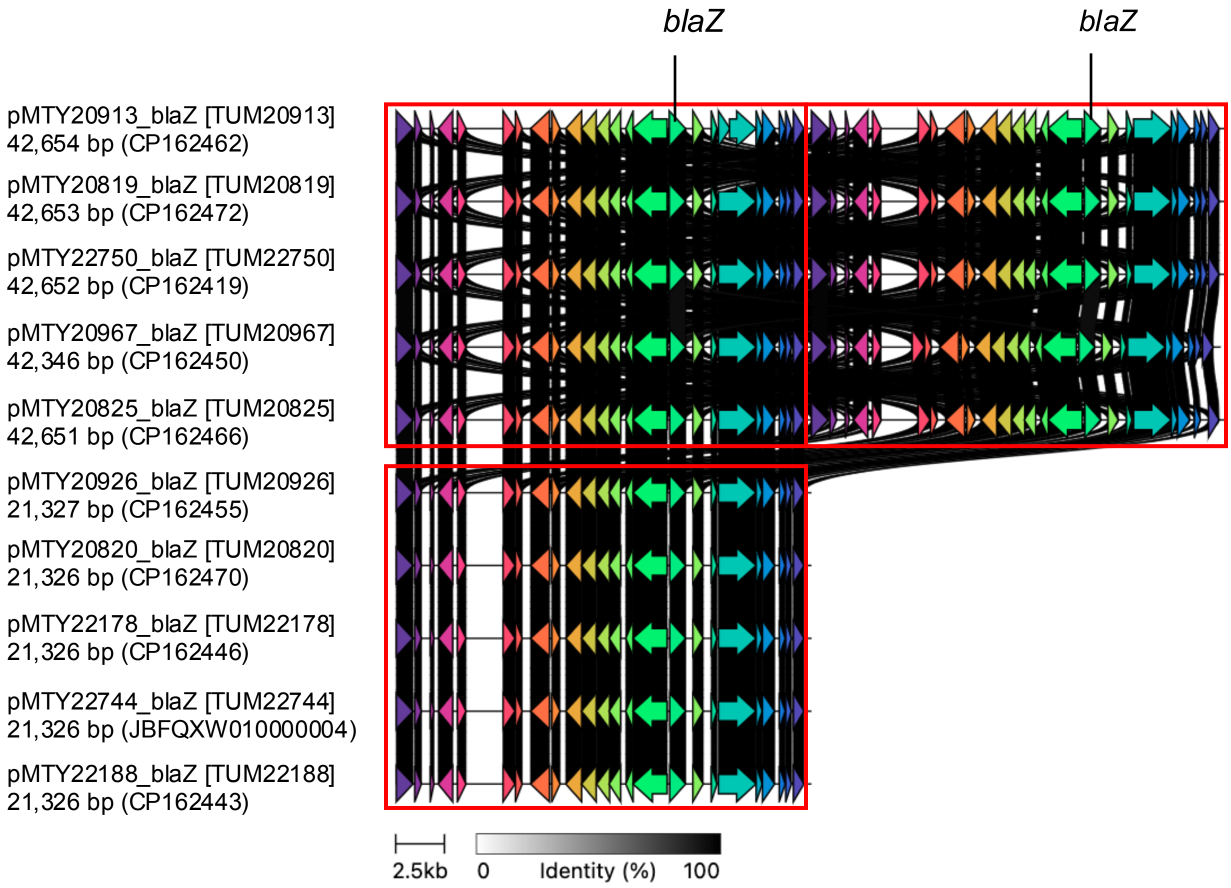

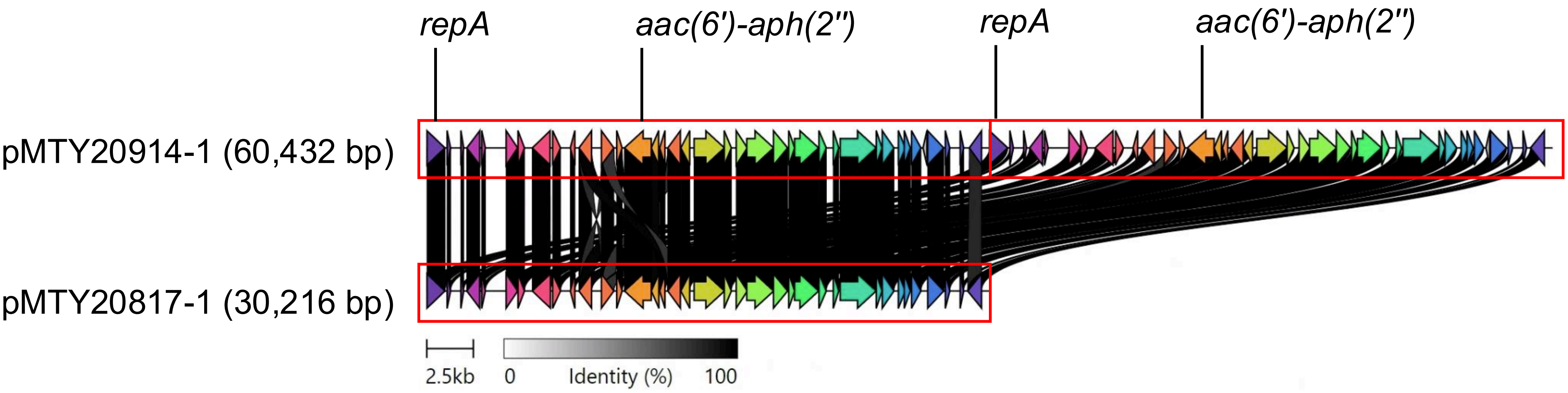

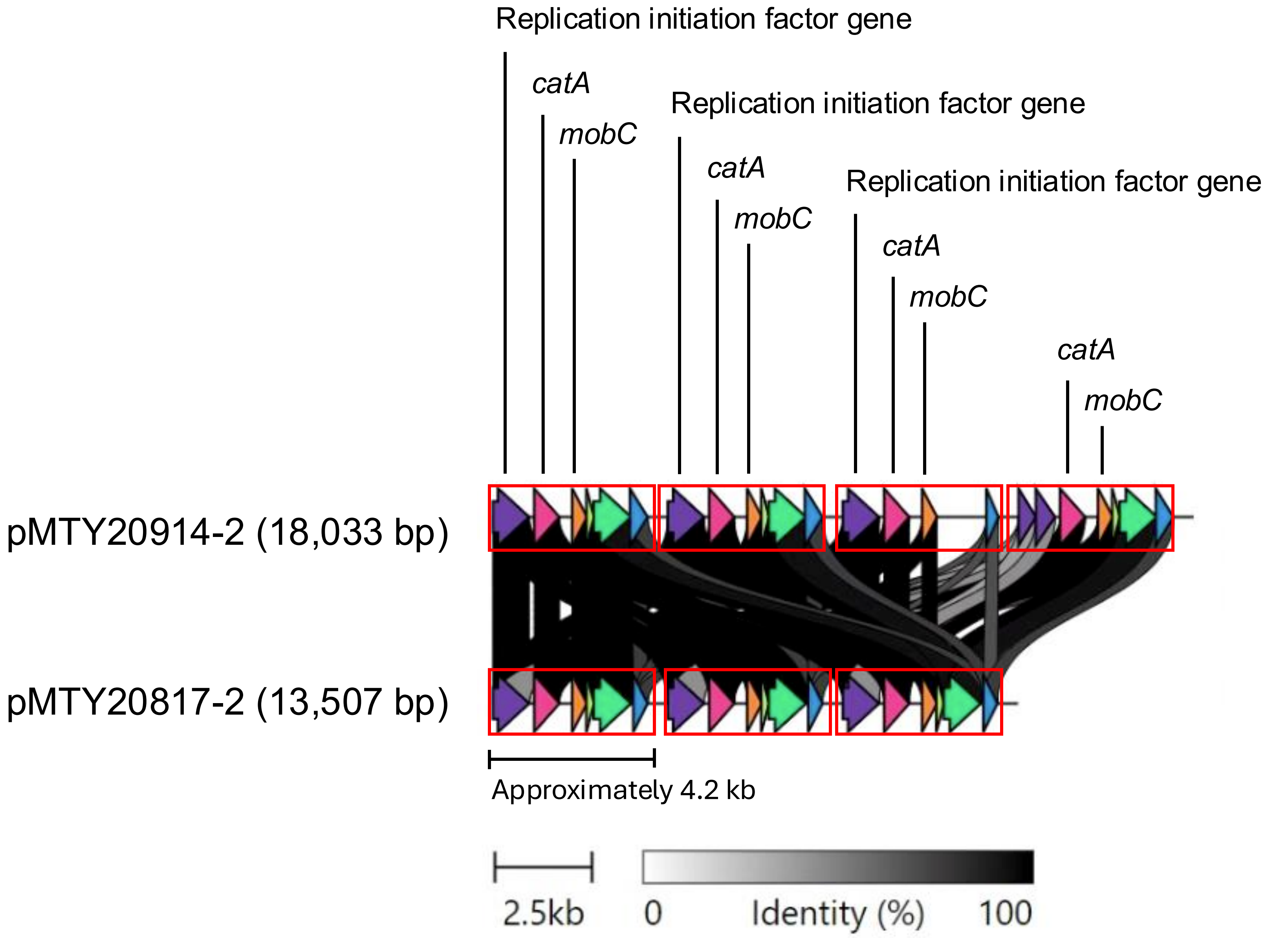

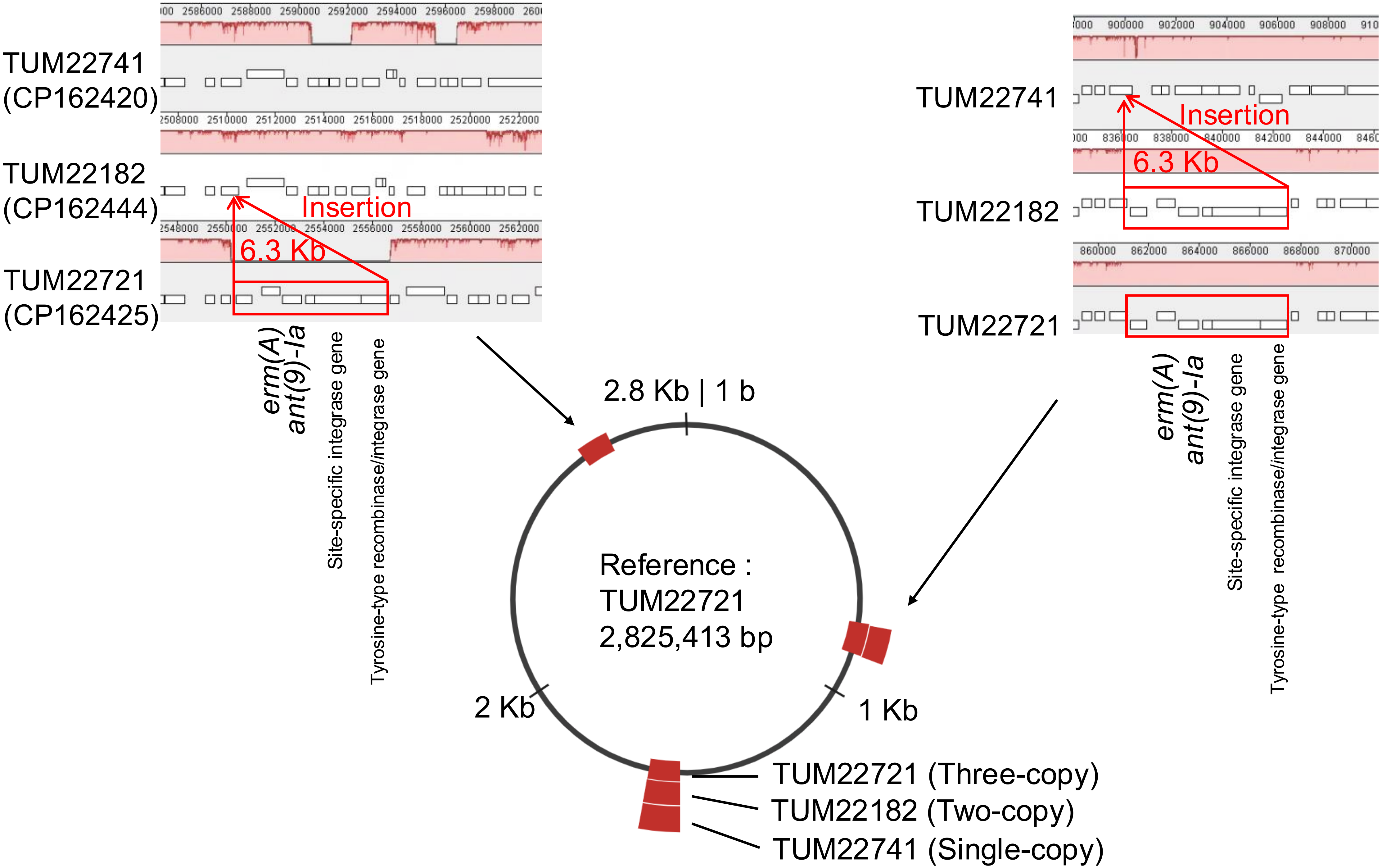
